# A randomized controlled trial of an interactive digital therapeutic for stress and burnout management

**DOI:** 10.1101/2025.10.13.25337485

**Authors:** Katharina M. Rischer, Linda T. Betz, Antje Riepenhausen, Björn Meyer, Gitta A. Jacob, Helge Frieling, Kamila Jauch-Chara

## Abstract

This pragmatic randomized controlled trial examined the effectiveness of *reviga*, a self-guided digital intervention based on cognitive behavioral therapy, in reducing work-related stress symptoms. A total of 290 adults experiencing significant stress and burnout were assigned to the intervention group (*reviga* + treatment as usual [TAU]; n = 147) or the control group (TAU only; n = 143). Intent-to-treat analyses showed that 3 months post-randomization, participants in the intervention group experienced significant positive effects on the primary outcome, perceived stress (Cohen’s *d* = 0.36), as well as on the secondary outcomes anxiety (*d* = 0.28), burnout (*d* = 0.31), occupational and social functioning (*d* = 0.31) and health-related quality of life (*d* = 0.35) compared to TAU. No effect was found for absenteeism quantified as the number of sick days. Effect sizes increased at 6-month follow-up. This study demonstrates that *reviga* represents a promising and scalable tool for workplace mental health support.

## Introduction

Work-related stress is a prevalent phenomenon across occupational sectors. A nationwide, representative survey by a major health insurer revealed that work is the top stressor in Germany, with 47% of respondents citing it as a source of stress ^1^. At the same time, many employees feel that their employers do not provide adequate resources for managing stress ^2^.

While short-term stress can potentially enhance task performance by facilitating adaptive responses to workplace demands, sustained exposure to stress has been associated with adverse effects on both physical and mental health ^3,4^. Chronic workplace stress is a significant risk factor for the development of common mental health disorders, including depression and anxiety, particularly in individuals experiencing effort-reward imbalances ^5^. Additionally, work-related stress has been implicated in sleep disturbances ^6,7^ and broader societal consequences, such as increased absenteeism ^8,9^, occupational disability ^10^, and constitutes a risk factor for physical diseases ^11^, contributing to a substantial public health burden. Given these wide-ranging implications, early intervention is essential to mitigate work-related stress and prevent burnout. As a key precursor to burnout, prolonged stress can lead to emotional, physical, and mental exhaustion, ultimately reducing professional efficacy and increasing detachment from work.

While traditional, face-to-face stress management interventions have demonstrated efficacy in reducing perceived stress levels ^12–14^, they are not readily available due to scarcity and long wait times for in-person resources, especially psychotherapy ^15,16^. At the same time, many patients are open to digital self-management interventions ^17^. However, 60% of those who self-manage express dissatisfaction with the outcomes of their efforts ^2^. Although numerous digital solutions for stress management are available, the vast majority lack an evidence base ^18^. Of those that are evidence-based, many are narrow in scope, typically focusing on specific techniques such as breathing exercises or relaxation techniques ^18,19^. This highlights a significant gap and underscores the urgent need for more accessible, evidence-based digital interventions that can provide timely, scalable support for work-related stress. A recent meta-analysis of 21 randomized controlled trials (RCTs) on web-based psychological interventions in the workplace found significant reductions in perceived stress, depression, and psychological distress (*d* = 0.37, 95% CI 0.23–0.50) ^20^. These findings demonstrate the potential of digital interventions to alleviate work-related psychological strain effectively.

*reviga* is a self-guided, digital intervention grounded in cognitive-behavioral therapy (CBT), specifically designed for individuals experiencing stress and burnout. As a flexible, resource-efficient solution, *reviga* requires no professional oversight and is accessible via the Internet at the user’s convenience. The program delivers personalized psychoeducation and therapeutic exercises tailored to individual needs and preferences. Features such as personalized content, automated reminders, and progress tracking are incorporated to promote consistent engagement and motivation.

This randomized controlled trial (RCT) evaluated the efficacy of *reviga* in alleviating perceived stress and burnout among adults experiencing work-related stress. The study included a Treatment as Usual (TAU) control group, which reflects the standard care typically available to individuals, including both informal workplace support programs and access to professional resources such as counseling or therapy. By evaluating the effectiveness of *reviga* within a diverse sample, this trial aimed to examine its utility for improving workplace mental health.

## Methods

### Recruitment and assessment

Prior to recruitment, the study “Evaluating the Effectiveness of a Digital Therapeutic (*reviga*) for people with stress and burnout - a randomized controlled trial (LAVENDER)” was registered in an international study register (NCT05998161). German-speaking participants were recruited through an online advertising campaign, primarily using targeted Google Ads to attract individuals experiencing stress and burnout. This approach was designed to efficiently reach the intended audience, directing interested individuals to a dedicated study website where they could find detailed information about the study and express their interest in participating. After electronically consenting to the study, participants completed an initial online survey. Following verification of the inclusion criteria, participants were randomly assigned to either the intervention or control group using an automated randomization system with a 1:1 allocation ratio (no blocked randomization, no stratification, concealed allocation, and without blinding of participants). Throughout the trial, participants in the intervention group were provided access to *reviga* in addition to TAU while those in the control group received only TAU and were given access to *reviga* once they completed the study. TAU allowed participants in both groups to continue or discontinue any treatments, such as psychotherapy or medication. Participants in both groups were asked to complete online assessments at three months (T1; primary assessment point for evaluating effectiveness) and again at six months (T2; follow-up). All study procedures were conducted online.

### Inclusion and exclusion criteria

Participants were eligible for inclusion if they were between 18 and 65 years of age, experienced above-average perceived stress levels (PSS score > 21) and burnout (OLBI ≥ 2.18). Additionally, they needed to be residing in Germany, be employed for at least 20 hours per week, and have maintained a stable treatment regimen for at least 30 days prior to enrollment. Participants were also required to provide informed consent to take part in the study. Individuals were excluded if they had plans to change their treatment within the next three months at the time of inclusion.

### Intervention

*reviga* is a self-guided online intervention based on cognitive-behavioral therapy (CBT) designed to support individuals experiencing stress and burnout. While the program has been described in previous publications ^21,22^, this is the first RCT evaluating its effectiveness. *reviga* combines psychoeducation, therapeutic exercises, and personalized techniques tailored to users’ needs (see **Table 1**). A key feature of *reviga* is its “simulated dialogue”, where users interact with predefined response options that dynamically shape their experience, a technique successfully used previously^23–26^. This approach personalizes content delivery while maintaining engagement. The program also includes simple homework assignments, flexible session pacing, and regular reminders to support adherence. Beyond its core dialogues, *reviga* offers supplementary tools such as guided audio exercises, PDF worksheets, and summary sheets. Users can opt to receive motivational messages via SMS or email, while self-monitoring questionnaires help track progress and behavioral changes over time.

**Table 1.**
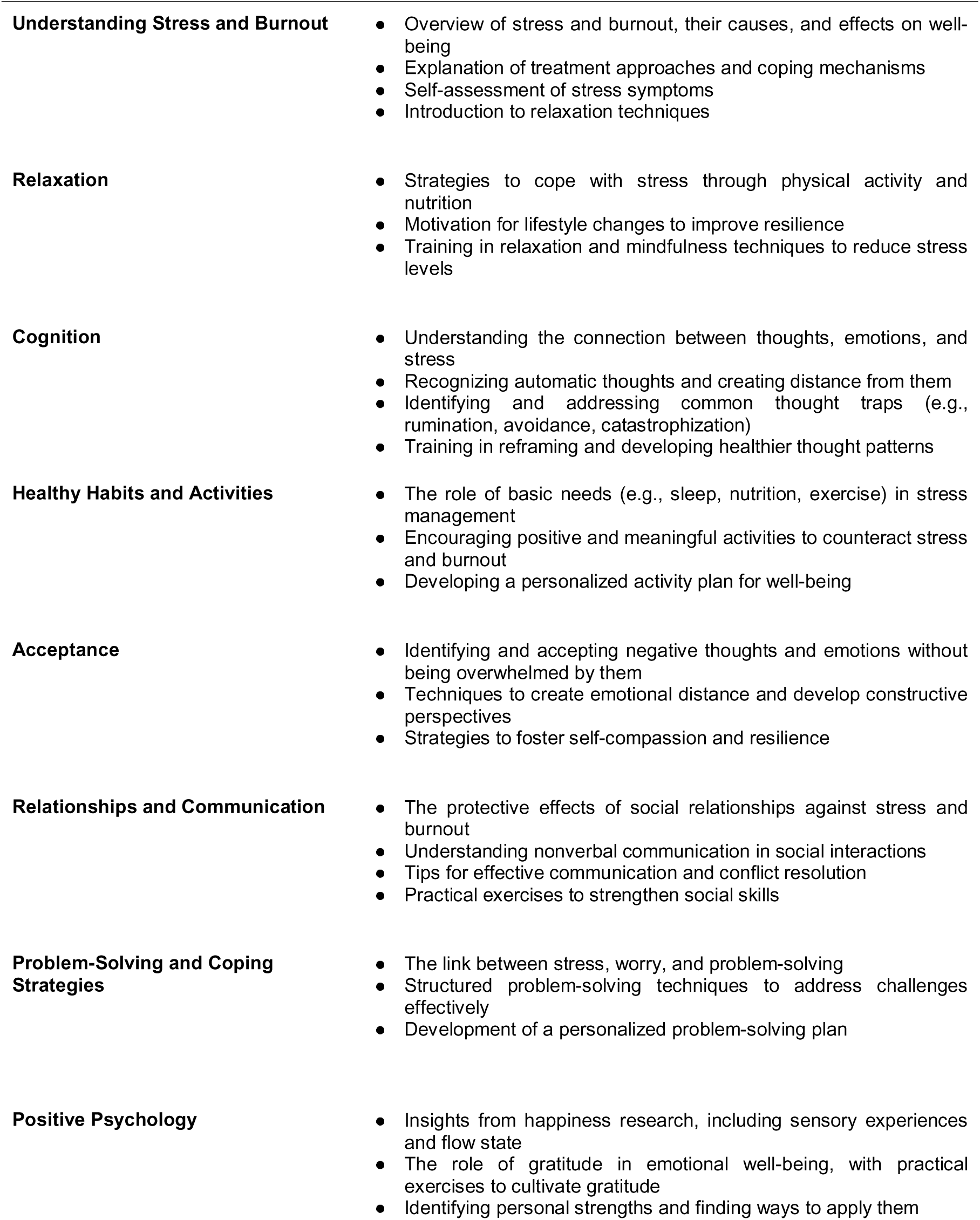
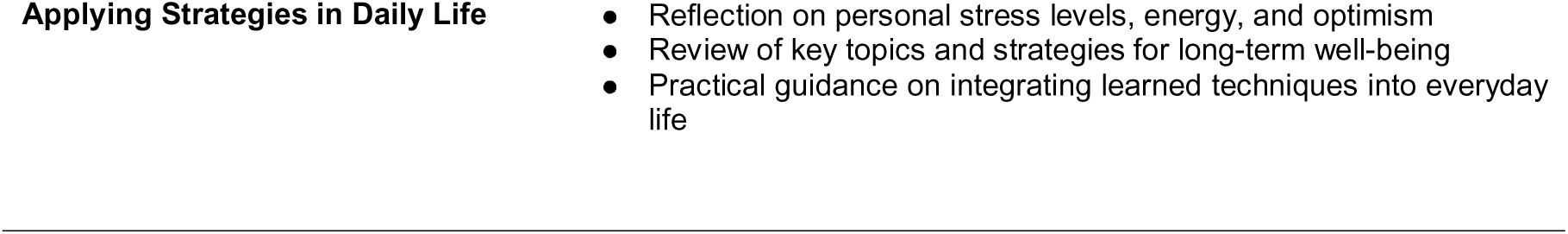
Content of *reviga*.

### Measures

Baseline data, as well as follow-up data at 3 months (T1) and 6 months (T2) post-randomization, were collected using a secure and encrypted online survey service (*Easyfeedback*). All data were obtained through self-report. Participants received email invitations to complete the assessments, and those who did not respond were followed up with up to three reminders and phone calls to encourage participation. The primary time point for assessing effectiveness was T1, with T2 serving as an additional time point to assess the durability of the effects.

### Primary Endpoint

The primary endpoint was the validated German version of the Perceived Stress Scale - 10 item version (PSS-10), a patient-reported outcome measure (PROM) ^27, 28^ at T1 (3 months post-randomization).

### Secondary Endpoints

Secondary outcomes included (1) anxiety symptoms, assessed with the Generalized Anxiety Disorder Assessment (GAD-7) ^29,30^; (2) functional impairment, measured by the Work and Social Adjustment Scale (WSAS) ^31,32^; (3) burnout, assessed with the Oldenburg Burnout Inventory (OLBI) ^33^; (4) health-related quality of life, measured by the Assessment of Quality of Life - 8 Dimensions (AQoL-8D) ^34,35^; and (5) absenteeism, assessed by the number of sick days in the past 90 days.

### Comorbidities and user satisfaction

The Web Screening Questionnaire (WSQ) was used to assess comorbidities in the sample ^36^. To assess satisfaction with *reviga,* participants were asked to rate how likely they were to recommend *reviga* to a friend or colleague on a scale from 0 (not likely at all) to 10 (extremely likely). Additionally, satisfaction was measured using the German version of the Client Satisfaction Questionnaire (CSQ-8)^37^.

### Statistical Analyses

We conducted intent-to-treat (ITT) analyses as the primary analysis for all outcomes, including all participants in the intervention and control groups, regardless of their intervention usage. By contrast, per-protocol (PP) analyses included only intervention participants who used *reviga* on at least two different days. Intervention effects at T1 were assessed using ANCOVA, adjusting for baseline values. Treatment effects are reported as baseline-adjusted mean differences with 95% CIs, and effect sizes were calculated using Cohen’s *d* based on estimated marginal means. To account for missing data, we employed multiple imputation using the *R* package *bootImpute* ^38^. As a sensitivity analysis, we applied the jump-to-reference (J2R) imputation method, a conservative approach that assumes intervention group participants who dropped out followed the trajectory of the control group, effectively treating dropout as a loss of any intervention benefits ^39,40^. ITT, J2R, and PP analyses were also repeated at T2 to evaluate the durability of effects. Additionally, a responder analysis classified participants as responders if they showed a ≥ 6-point improvement at T1 (corresponding to 15% of the scale range ^41^), and proportions of responders were compared using χ² tests. To control for multiple comparisons, we applied a gatekeeping strategy, testing secondary endpoints sequentially ^42^.

### Sample Size

An a priori power analysis, based on an anticipated effect size of *d* = 0.37 ^20^, with a power of 0.80 at an α = 0.05 resulted in a required minimum sample size of N = 232, n = 116 per arm. Considering an expected drop-out rate of 20% of participants, the minimum sample size was set a priori at N = 290 (n = 145 per arm).

## Results

### Description of trial participants

Participants were recruited between September 2023 and November 2023 (follow-up period up to June 2024) via online advertisement. 936 people were screened for participation. Of these, 290 met all specified inclusion criteria and were randomized to the intervention (n = 147) und control group (n = 143). Participant characteristics at baseline are presented in **Table 2**. Participants were in their mid-40s on average, mostly female (86.2%) and well-educated (60.3% university graduates). About 60% were employed full-time, while the rest worked part-time. Sick leave varied, with about a third reporting no days off in the past month, while a similar proportion had more than 10 days of sick leave. Mental health screening showed high rates of positive results for generalized anxiety (90.7%), post-traumatic stress disorder (61.7%), specific phobia (59.3%), and depression (47.2%). Importantly however, the WSQ overestimates the true prevalence of diagnoses and should not be interpreted as a clinical assessment ^43^. Psychotherapy history was reported by about two-thirds of participants, and around one in ten were currently taking antidepressants. Reasons for exclusion, as well as attrition and reasons for drop-out are provided in the study flow chart (**Figure 1**).

**Figure 1.**
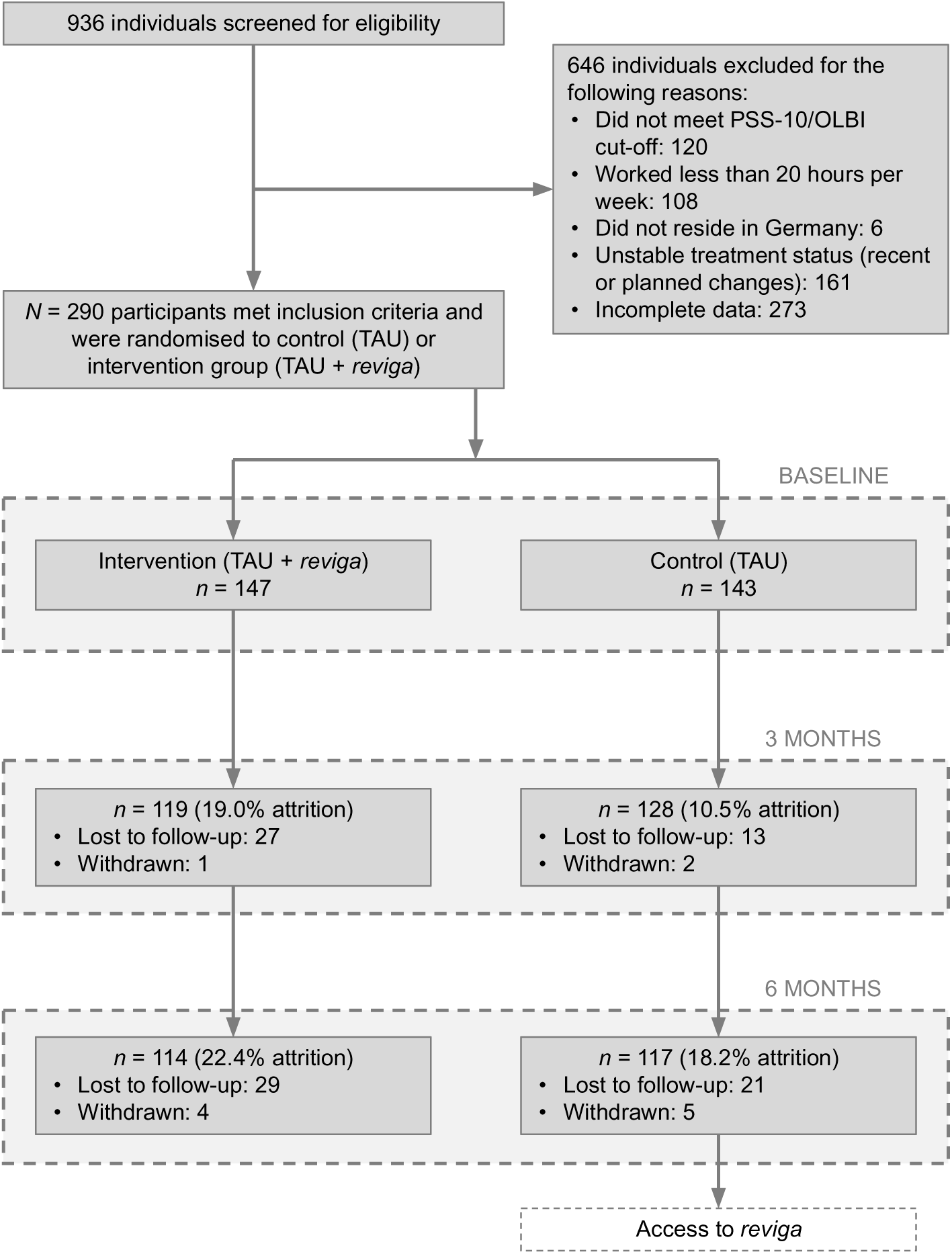
Participant Flow Chart. *Note:* TAU = treatment as usual.

**Table 2.**
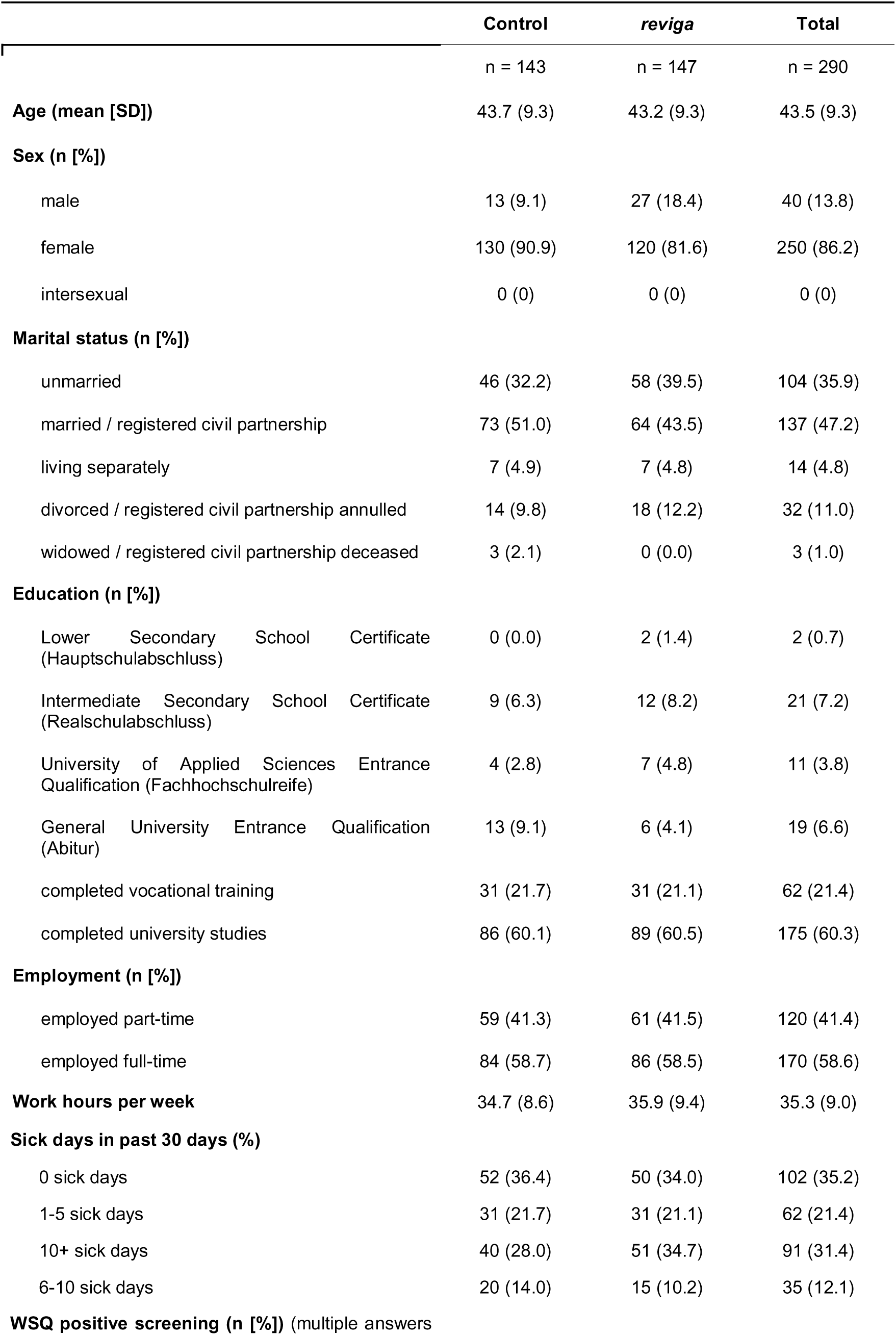
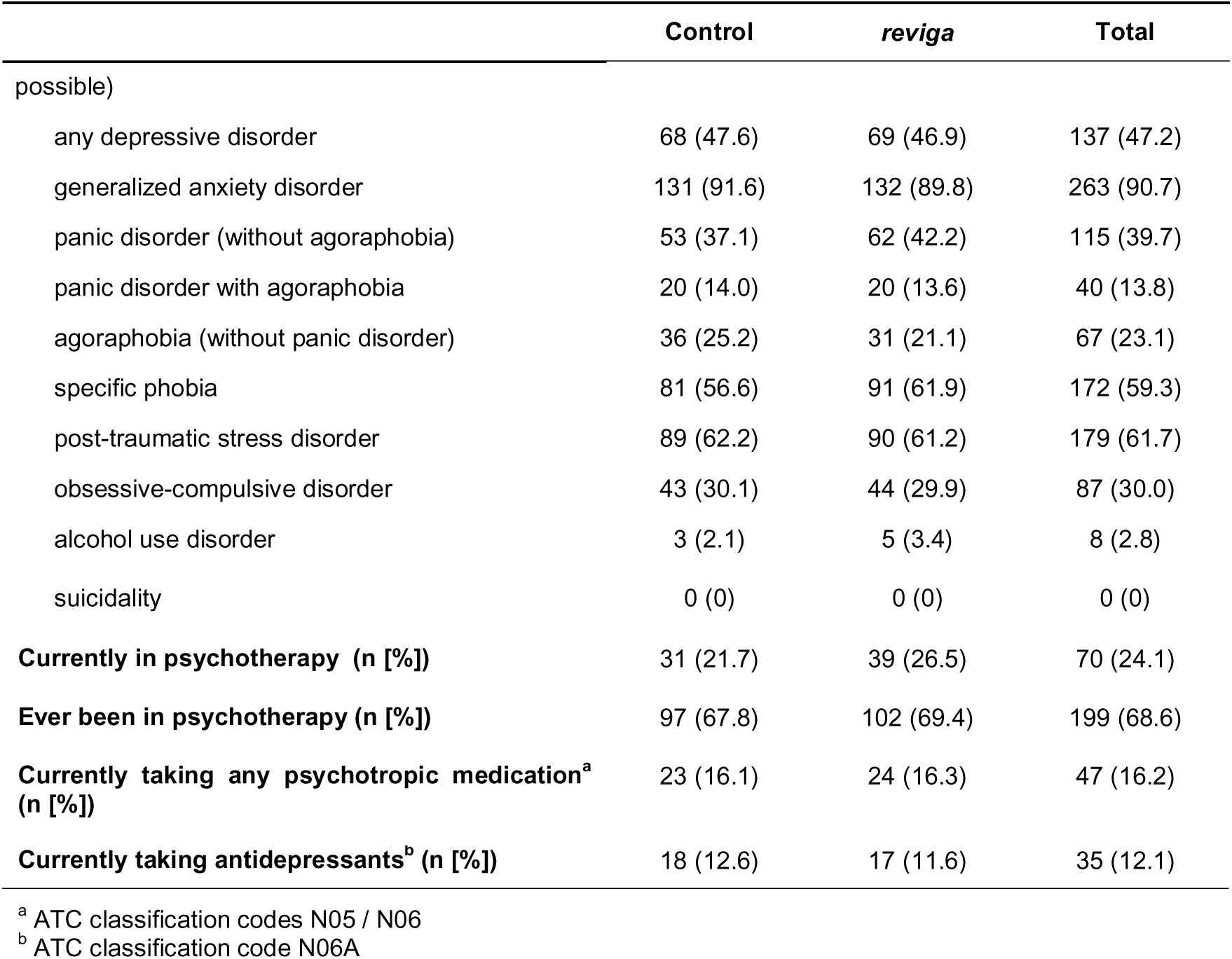
Participant demographics and clinical characteristics at baseline.

|  | Control | *reviga* | Total |
| --- | --- | --- | --- |
|  | n = 143 | n = 147 | n = 290 |
| **Age (mean [SD])** | 43.7 (9.3) | 43.2 (9.3) | 43.5 (9.3) |
| **Sex (n [%])** |  |  |  |
| male | 13 (9.1) | 27 (18.4) | 40 (13.8) |
| female | 130 (90.9) | 120 (81.6) | 250 (86.2) |
| intersexual | 0 (0) | 0 (0) | 0 (0) |
| **Marital status (n [%])** |  |  |  |
| unmarried | 46 (32.2) | 58 (39.5) | 104 (35.9) |
| married / registered civil partnership | 73 (51.0) | 64 (43.5) | 137 (47.2) |
| living separately | 7 (4.9) | 7 (4.8) | 14 (4.8) |
| divorced / registered civil partnership annulled | 14 (9.8) | 18 (12.2) | 32 (11.0) |
| widowed / registered civil partnership deceased | 3 (2.1) | 0 (0.0) | 3 (1.0) |
| **Education (n [%])** |  |  |  |
| Lower Secondary School Certificate (Hauptschulabschluss) | 0 (0.0) | 2 (1.4) | 2 (0.7) |
| Intermediate Secondary School Certificate (Realschulabschluss) | 9 (6.3) | 12 (8.2) | 21 (7.2) |
| University of Applied Sciences Entrance Qualification (Fachhochschulreife) | 4 (2.8) | 7 (4.8) | 11 (3.8) |
| General University Entrance Qualification (Abitur) | 13 (9.1) | 6 (4.1) | 19 (6.6) |
| completed vocational training | 31 (21.7) | 31 (21.1) | 62 (21.4) |
| completed university studies | 86 (60.1) | 89 (60.5) | 175 (60.3) |
| **Employment (n [%])** |  |  |  |
| employed part-time | 59 (41.3) | 61 (41.5) | 120 (41.4) |
| employed full-time | 84 (58.7) | 86 (58.5) | 170 (58.6) |
| **Work hours per week** | 34.7 (8.6) | 35.9 (9.4) | 35.3 (9.0) |
| **Sick days in past 30 days (%)** |  |  |  |
| 0 sick days | 52 (36.4) | 50 (34.0) | 102 (35.2) |
| 1-5 sick days | 31 (21.7) | 31 (21.1) | 62 (21.4) |
| 10+ sick days | 40 (28.0) | 51 (34.7) | 91 (31.4) |
| 6-10 sick days | 20 (14.0) | 15 (10.2) | 35 (12.1) |
| **WSQ positive screening (n [%])** (multiple answers |  |  |  |

|  | **Control** | **reviga** | **Total** |
| --- | --- | --- | --- |
| possible) |  |  |  |
| any depressive disorder | 68 (47.6) | 69 (46.9) | 137 (47.2) |
| generalized anxiety disorder | 131 (91.6) | 132 (89.8) | 263 (90.7) |
| panic disorder (without agoraphobia) | 53 (37.1) | 62 (42.2) | 115 (39.7) |
| panic disorder with agoraphobia | 20 (14.0) | 20 (13.6) | 40 (13.8) |
| agoraphobia (without panic disorder) | 36 (25.2) | 31 (21.1) | 67 (23.1) |
| specific phobia | 81 (56.6) | 91 (61.9) | 172 (59.3) |
| post-traumatic stress disorder | 89 (62.2) | 90 (61.2) | 179 (61.7) |
| obsessive-compulsive disorder | 43 (30.1) | 44 (29.9) | 87 (30.0) |
| alcohol use disorder | 3 (2.1) | 5 (3.4) | 8 (2.8) |
| suicidality | 0 (0) | 0 (0) | 0 (0) |
| **Currently in psychotherapy (n [%])** | 31 (21.7) | 39 (26.5) | 70 (24.1) |
| **Ever been in psychotherapy (n [%])** | 97 (67.8) | 102 (69.4) | 199 (68.6) |
| **Currently taking any psychotropic medication<sup>a</sup> (n [%])** | 23 (16.1) | 24 (16.3) | 47 (16.2) |
| **Currently taking antidepressants<sup>b</sup> (n [%])** | 18 (12.6) | 17 (11.6) | 35 (12.1) |

<sup>a</sup> ATC classification codes N05 / N06
<sup>b</sup> ATC classification code N06A

### Study outcomes

ITT analyses demonstrated significant intervention effects of small magnitude across all endpoints, except on absenteeism, at three and six months (see **Table 3** and **Figure 2**). Engagement with *reviga* was high, with 95.2% of participants in the intervention group (140 of 147) registering for use. On average, they interacted with the program on 17 different days (SD = 18.0), and 87.8% (129 of 147) met the predefined minimum usage threshold of at least two days. As a result, the PP dataset included 272 participants (129 from the intervention group and all 143 controls). PP analyses indicated slightly stronger intervention effects (see **Table 4**), and a conservative sensitivity analysis using J2R imputation consistently supported all findings (see **supplementary Table 1**).

**Figure 2.**
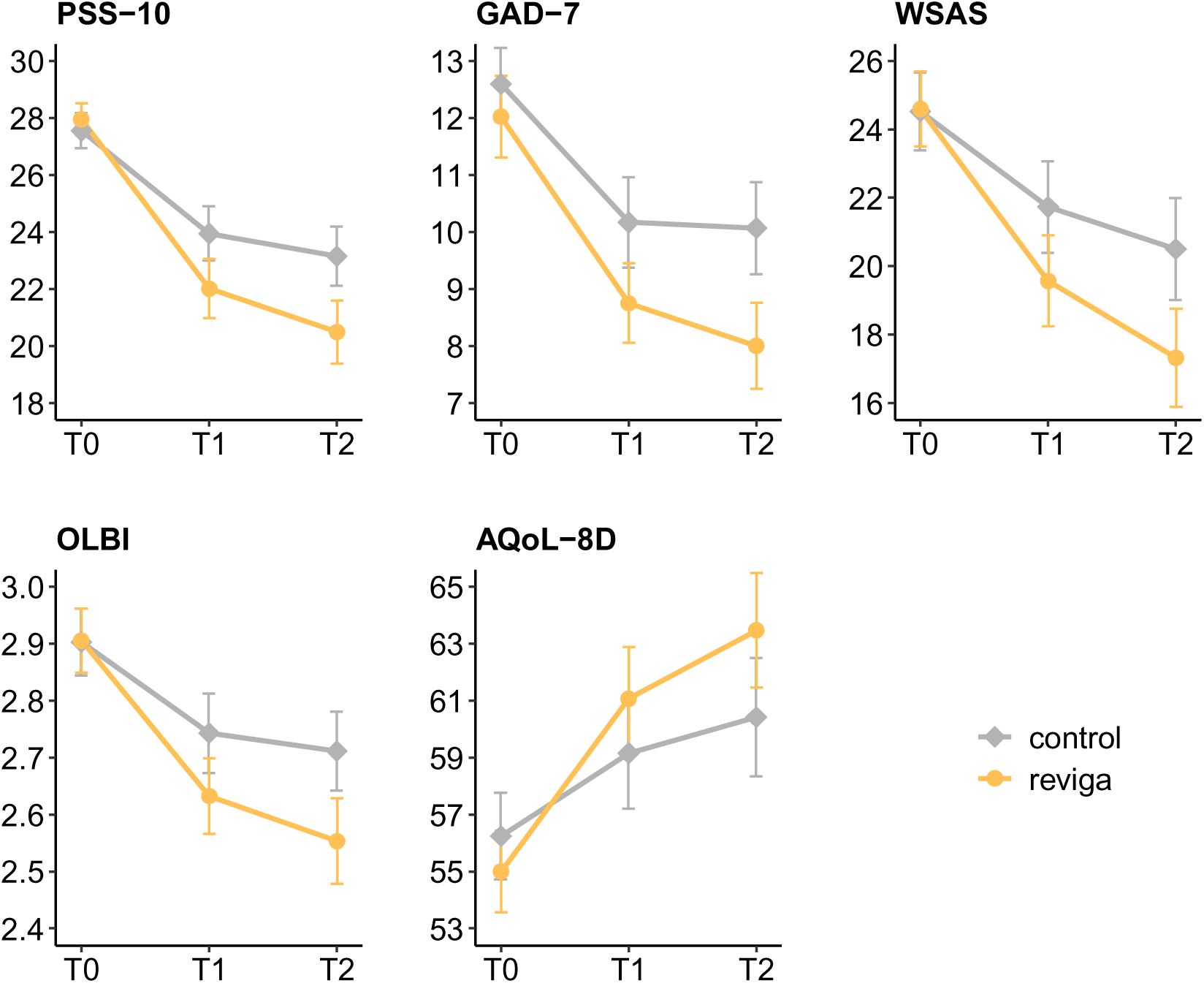
Mean scores for primary and secondary endpoints for the *reviga* group and control group across the study period, derived from the intention-to-treat (ITT) analysis. Error bars represent the 95% Confidence Interval (CI). Time points are T0 = Baseline, T1 = 3 months, T2 = 6 months. *Note:* AQoL-8D = Assessment of Quality of Life - 8 Dimensions; GAD-7 = Generalized Anxiety Disorder Assessment; OLBI = Oldenburg Burnout Inventory; PSS-10 = Perceived Stress Scale - 10 items; WSAS = Work and Social Adjustment Scale.

**Table 3.** Results of primary and secondary endpoints for ITT analyses.

|  | Time | Control |  |  | reviga |  |  | ANCOVA |  |  |  |
| --- | --- | --- | --- | --- | --- | --- | --- | --- | --- | --- | --- |
| | | n | mean | SD | n | mean | SD | Treatment effect<sup>a</sup><br>(95% CI) | p-value | Partial $\eta^2$ | Cohen's *d*<br>(95% CI)<sup>b</sup> |
| PSS-10 | T0 | 143 | 27.6 | 3.7 | 147 | 28.0 | 3.5 | - | - | - | - |
|  | T1 | 143 | 23.9 | 5.8 | 147 | 22.0 | 6.4 | -2.1<br>(-3.6, -0.7) | 0.005 | 0.04 | 0.36<br>(0.12, 0.61) |
|  | T2 | 143 | 23.2 | 6.3 | 147 | 20.5 | 6.8 | -2.9<br>(-4.6, -1.2) | < .001 | 0.05 | 0.46<br>(0.18, 0.74) |
| GAD-7 | T0 | 143 | 12.6 | 3.9 | 147 | 12.0 | 4.4 | - | - | - | - |
|  | T1 | 143 | 10.2 | 4.9 | 147 | 8.7 | 4.3 | -1.2<br>(-2.2, -0.1) | 0.031 | 0.02 | 0.28<br>(0.02, 0.53) |
|  | T2 | 143 | 10.1 | 4.9 | 147 | 8.0 | 4.7 | -1.8<br>(-3, -0.7) | 0.002 | 0.04 | 0.41<br>(0.15, 0.67) |
| WSAS | T0 | 143 | 24.5 | 7.0 | 147 | 24.6 | 6.8 | - | - | - | - |
|  | T1 | 143 | 21.7 | 8.2 | 147 | 19.6 | 8.2 | -2.2<br>(-4.1, -0.3) | 0.024 | 0.03 | 0.31<br>(0.04, 0.57) |
|  | T2 | 143 | 20.5 | 9.1 | 147 | 17.3 | 8.9 | -3.2<br>(-5.3, -1.1) | 0.002 | 0.04 | 0.39<br>(0.14, 0.65) |
| OLBI | T0 | 143 | 2.9 | 0.4 | 147 | 2.9 | 0.3 | - | - | - | - |
|  | T1 | 143 | 2.7 | 0.4 | 147 | 2.6 | 0.4 | -0.1<br>(-0.2, 0) | 0.018 | 0.03 | 0.31<br>(0.06, 0.55) |
|  | T2 | 143 | 2.7 | 0.4 | 147 | 2.6 | 0.5 | -0.2<br>(-0.3, -0.1) | 0.003 | 0.04 | 0.40<br>(0.13, 0.66) |
| AQoL-8D | T0 | 143 | 56.2 | 9.3 | 147 | 55.0 | 8.9 | - | - | - | - |
|  | T1 | 143 | 59.2 | 11.9 | 147 | 61.1 | 11.3 | 3.0<br>(0.7, 5.3) | 0.01 | 0.03 | 0.35<br>(0.09, 0.62) |
|  | T2 | 143 | 60.4 | 12.7 | 147 | 63.5 | 12.4 | 4.1<br>(1.6, 6.6) | 0.002 | 0.05 | 0.42<br>(0.16, 0.68) |
| Sick days | T0 | 143 | 12.1 | 21.1 | 147 | 14.3 | 23.9 | - | - | - | - |
|  | T1 | 143 | 15.5 | 26.1 | 147 | 17.5 | 26.9 | 0.4<br>(-4.5, 5.2) | 0.883 | 0 | -0.02<br>(-0.26, 0.23) |
|  | T2 | 143 | 13.0 | 24.5 | 147 | 12.4 | 23.8 | -1.9<br>(-7, 3.2) | 0.461 | 0.01 | 0.09<br>(-0.16, 0.35) |

<sup>a</sup> Group difference on original scale 3 months (T1) and 6 months (T2) after baseline, adjusted for baseline scores.<sup>b</sup> Based on baseline-adjusted means; positive values show effects in favor of the intervention group.

**Table 4.** Results of primary and secondary endpoints for PP analyses.

|  | Time | Control |  |  | reviga |  |  | ANCOVA |  |  |  |
| --- | --- | --- | --- | --- | --- | --- | --- | --- | --- | --- | --- |
| | | n | mean | SD | n | mean | SD | Treatment effect<sup>a</sup><br>(95% CI) | p-value | Partial $\eta^2$ | Cohen's *d*<br>(95% CI)<sup>b</sup> |
| PSS-10 | T0 | 143 | 27.6 | 3.7 | 129 | 28.0 | 3.7 | - | - | - | - |
|  | T1 | 143 | 24.0 | 5.8 | 129 | 22.0 | 6.6 | -2.2<br>(-3.8, -0.6) | 0.006 | 0.04 | 0.38<br>(0.11, 0.65) |
|  | T2 | 143 | 23.2 | 6.3 | 129 | 20.1 | 6.8 | -3.4<br>(-5, -1.8) | < .001 | 0.07 | 0.54<br>(0.27, 0.81) |
| GAD-7 | T0 | 143 | 12.6 | 3.9 | 129 | 12.0 | 4.5 | - | - | - | - |
|  | T1 | 143 | 10.2 | 4.9 | 129 | 8.6 | 4.3 | -1.3<br>(-2.5, -0.2) | 0.017 | 0.03 | 0.32<br>(0.06, 0.59) |
|  | T2 | 143 | 10.1 | 4.9 | 129 | 7.9 | 4.8 | -1.9<br>(-3.1, -0.8) | 0.001 | 0.05 | 0.43<br>(0.17, 0.70) |
| WSAS | T0 | 143 | 24.5 | 7.0 | 129 | 24.8 | 6.7 | - | - | - | - |
|  | T1 | 143 | 21.7 | 8.2 | 129 | 19.4 | 8.1 | -2.4<br>(-4.3, -0.5) | 0.011 | 0.03 | 0.34<br>(0.08, 0.60) |
|  | T2 | 143 | 20.6 | 9.1 | 129 | 16.9 | 8.7 | -3.8<br>(-5.9, -1.8) | < .001 | 0.06 | 0.47<br>(0.22, 0.73) |
| OLBI | T0 | 143 | 2.9 | 0.4 | 129 | 2.9 | 0.3 | - | - | - | - |
|  | T1 | 143 | 2.7 | 0.4 | 129 | 2.6 | 0.4 | -0.1<br>(-0.2, 0) | 0.02 | 0.03 | 0.31<br>(0.05, 0.56) |
|  | T2 | 143 | 2.7 | 0.4 | 129 | 2.5 | 0.5 | -0.2<br>(-0.3, -0.1) | < .001 | 0.05 | 0.46<br>(0.19, 0.72) |
| AQoL-8D | T0 | 143 | 56.2 | 9.3 | 129 | 54.6 | 9.0 | - | - | - | - |
|  | T1 | 143 | 59.2 | 11.9 | 129 | 61.3 | 11.3 | 3.5<br>(1.2, 5.7) | 0.003 | 0.04 | 0.41<br>(0.14, 0.67) |
|  | T2 | 143 | 60.4 | 12.7 | 129 | 63.8 | 12.3 | 4.8<br>(2.2, 7.5) | < .001 | 0.06 | 0.50<br>(0.22, 0.79) |
| Sick days | T0 | 143 | 12.2 | 21.2 | 129 | 14.5 | 24.0 | - | - | - | - |
|  | T1 | 143 | 15.6 | 26.1 | 129 | 17.5 | 26.9 | 0.1<br>(-5.2, 5.4) | 0.959 | 0.01 | -0.01<br>(-0.27, 0.26) |
|  | T2 | 143 | 13.1 | 24.7 | 129 | 12.8 | 24.3 | -1.7<br>(-6.9, 3.5) | 0.532 | 0.01 | 0.08<br>(-0.17, 0.33) |

<sup>a</sup> Group difference on original scale 3 months (T1) and 6 months (T2) after baseline, adjusted for baseline scores.<sup>b</sup> Based on baseline-adjusted means; positive values show effects in favor of the intervention group.

Responder analysis based on complete cases further suggested a higher rate of clinically relevant improvements (defined as ≥6 points on the PSS) in the intervention group than in the control group, though the difference did not reach statistical significance. At T1, 42.9% (51 of 119) of intervention participants were classified as responders compared to 35.9% (46 of 128) in the control group (χ² = 1.24, *p* = .266; OR = 1.34, 95% CI [0.80, 2.23]).

### Adverse effects and user satisfaction

No adverse events related to the use of *reviga* or any adverse device effects were observed. The average score on the NPS was 6.7 (SD = 2.7) at T1 and 6.5 (SD = 3.0) at T2, suggesting that, overall, participants were more likely to recommend *reviga* to a friend or colleague than not. Mean score on the CSQ-8 was 19.8 (SD = 1.3) at T1 and 19.7 (SD = 1.7) at T2.

## Discussion

The LAVENDER-RCT aimed to assess the effectiveness of *reviga*, a self-guided digital intervention based on CBT among individuals experiencing work-related stress. The findings indicate that *reviga* leads to a significant reduction in stress-related symptoms and an improvement in functioning and health-related QoL after 3 months (T1), with these effects increasing in magnitude after 6 months (T2). No significant intervention effects were observed on sick days. Results were confirmed and remained consistent in a conservative sensitivity analysis, which assumed that any dropout in the intervention group would result in a loss of intervention effects. Pre-specified PP analyses suggested stronger intervention effects in participants who used *reviga* beyond initial exposure (i.e., on at least two different days). Given the prevalence of work-related stress and limited employer support, these findings highlight *reviga* as a scalable option for individuals with limited access to in-person resources.

With a standardized effect size of *d* = 0.36 on perceived stress after three months, *reviga* performs in line with the *d* = 0.37 average effect size reported in a meta-analysis of digital interventions for employee well-being ^20^, supporting its expected effectiveness and reinforcing the validity of the results. Importantly, data suggest that *reviga* outperforms other CBT-based approaches, which show a lower average effect size of *d* = 0.25 ^20^. This suggests that *reviga* may offer a particularly effective application of relevant CBT principles in a digital format. A contributing factor could be its integration of key features identified as enhancing effectiveness and engagement for digital interventions for stress and burnout, such as persuasive system design features like tailoring and self-monitoring and secondary delivery modalities (e.g., email and text messages) ^20^. Interestingly, *reviga* demonstrated increasing effects on all outcomes over time, with effect sizes on the primary endpoint, perceived stress, reaching *d* = 0.46 after 6 months. Work-related behaviors, including stress responses and coping mechanisms, are developed over long periods and may take time to unlearn, particularly when they have become habitual or reinforced by workplace dynamics. Thus, as users repeatedly engage with therapeutic content, refine their decision-making patterns, and apply newly acquired skills more confidently in their routines, improvements may become more pronounced over time. Such a trajectory aligns with theories of cognitive and behavioral adaptation, which propose that sustained engagement is necessary for new strategies to be ingrained and yield meaningful change ^44^. Consistently, PP analyses revealed stronger effects (up to *d* = 0.54 for perceived stress at T2), suggesting that participants who engaged more actively with the intervention experienced greater benefits.

*reviga* had broad positive effects on the sequelae of chronic stress, including significant reductions in burnout and anxiety as well as improvements in functioning and health-related quality of life. However, it did not show an effect on sick days. This may be due to the fact that more than half of participants had minimal or no sick days at baseline, reducing the potential to detect an intervention effect. Absenteeism is also a lagging indicator, suggesting that longer follow-up may be needed to assess whether use of *reviga* may lead to fewer sick days in the long term.

Overall, the intervention effects observed for *reviga* are comparable with in-person occupational interventions in terms of impact on psychosocial outcomes ^20^. At the same time, the improvements observed in the control group suggest TAU also contributed to stress reduction, potentially reflecting growing awareness and accessibility of mental health resources in the workplace in Germany ^45^. Moreover, the experience of study participation itself, with its structured assessments, reflection prompts, and increased attention to stress-related symptoms, may have facilitated improvements ^46^. While between-group differences at T1 were therefore modest, the increase in *reviga*’s effects over time suggests that it may provide additional long-term benefits beyond TAU as users become more adept in applying their knowledge and acquired skills in their work routines, as discussed above.

While the intervention effects of *reviga* are promising, there is potential to enhance user satisfaction, especially when compared to structurally similar interventions recently developed and evaluated by the same developer in other indications ^23–25^. One possible explanation may be that individuals experiencing work-related stress and burnout often develop cognitive biases associated with heightened negativity ^47,48^, which may have influenced their perception of the intervention. Thus, future research should examine ways to optimize *reviga* for different user profiles. Research suggests that human guidance in digital interventions may be an important factor to enhance engagement and effectiveness in general ^49,50^ and in people with burnout specifically ^20,51^; however, use of human resources must be carefully balanced with the need for scalability. As a complementary approach, personalization supported by large language models could help provide guidance in digital interventions within the boundaries of evidence-based and responsible implementation ^52^. Determining the most effective way to implement such solutions remains an important task for future research.

The present RCT has several key strengths. First, with a total sample size of N = 290, this trial is among the largest conducted to date on digital interventions for work-related stress and burnout, significantly enhancing the validity and generalizability of its findings ^20^. Second, the study adopted a pragmatic design, ensuring that it closely mirrors real-world conditions. Its broad inclusion and exclusion criteria capture the diverse experiences of individuals facing work-related stress and burnout, further strengthening the study’s relevance and impact. Third, the trial incorporated follow-up assessments at two time points (3 and 6 months post-randomization), allowing for a comprehensive evaluation of both immediate and sustained intervention effects.

We also acknowledge several limitations. First, participant dropout remains a relevant factor. While the present dropout rates are within an expected range and similar to those observed in previous trials of structurally comparable interventions ^23–26^, understanding reasons for dropout, such as perceived burden, motivation, or user expectations, could provide insights for further optimizing the intervention. Second, while the results at T1 were statistically significant for the primary endpoint, they did not meet the pre-defined threshold for clinical relevance. In the absence of a validated minimally important clinical difference, we had to rely on a generic, distribution-based criterion to define response ^41^. However, this may not be the most appropriate cut-off for identifying clinically meaningful change in perceived stress. Additionally, the primary endpoint assessed general rather than work-related stress, which may have been too unspecific to show clinically relevant changes after only 3 months. Third, our sample predominantly consisted of women. This overrepresentation may be partially explained by sex differences in the experience of burnout ^53,54^. Additionally, women are more likely to seek help for mental health concerns, including utilizing psychotherapy and other specialized services ^55,56^. This trend also extends to digital interventions, where most users are women ^57,58^. As such, further data could strengthen our understanding of *reviga*’s effectiveness for men. Fourth, digital interventions for stress and burnout show promise but may not be universally applicable. Individuals hesitant to engage with digital platforms may be beyond their reach and were likely underrepresented due to online recruitment. Consequently, findings may primarily generalize to those equipped for and motivated to use digital interventions rather than the broader population experiencing burnout that could benefit from CBT-based interventions ^23^.

In summary, *reviga* demonstrates consistent effectiveness in reducing stress, anxiety, and burnout symptoms while improving functioning and health-related quality of life, with effects increasing over six months. Notably, as a fully self-guided intervention, *reviga* requires no additional human resources, making it a highly scalable solution. PP analyses further support its effectiveness, showing stronger effects among participants who engaged beyond initial exposure. These findings contribute to the growing evidence base supporting digital self-help interventions as an effective and accessible approach for managing work-related stress and burnout. Given the increasing demand for evidence-based digital mental health treatments and the limited availability of in-person stress management programs, *reviga* has the potential to address a critical gap in workplace mental health care.

## Data Availability

All data produced in the present study are available upon reasonable request to the authors.

## Acknowledgements

This trial was funded by GAIA (Hamburg, Germany). The authors thank the individuals who participated in the *reviga* trial.

