## Supplementary Materials for "A randomized controlled trial of an interactive digital therapeutic for stress and burnout management"

### Supplementary Methods

All analyses were conducted using *R*, version 4.4.1 (R Core Team, 2024).

Intervention effects of numerical outcomes at T1 were assessed using ANCOVA, with the respective outcome at T1 as the dependent variable, treatment condition (intervention vs. control) as the independent variable, and baseline values as a covariate. Treatment effects, including baseline-adjusted mean group differences and corresponding 95% CIs, are reported on the original scale, with significance determined by the corresponding p-value. Between-group effect sizes were calculated using Cohen's *d* based on the baseline-adjusted means from the ANCOVA model (estimated marginal means) with the *R* package *emmeans* [1].

The primary analysis was conducted as an intent-to-treat (ITT) analysis, including all randomized participants, regardless of intervention usage. Missing data at T1 were imputed using multiple imputation based on baseline values and sociodemographic and clinical variables (age, sex, weekly work hours, psychotherapy at baseline, and antidepressant use). Specifically, we applied von Hippel's bootstrapped maximum likelihood multiple imputation method [2], which improves standard multiple imputation by incorporating an explicit bootstrap step to better approximate the sampling distribution of missing data. The imputation process was performed using the *bootImpute* and *mice* packages in *R* [3].

As a sensitivity analysis, we applied reference-based multiple imputation using the conservative jump-to-reference (J2R) approach, which assumes that participants in the intervention group who dropped out would follow the trajectory of the control group, effectively treating dropout as a loss of intervention benefits [4,5]. By making this conservative assumption, J2R provides a worst-case scenario for treatment effects, ensuring that observed benefits are not driven by selective dropout. J2R was performed using the *R* package *bootImpute*.

Lastly, pre-specified per-protocol (PP) analyses were conducted, including only intervention group participants who engaged with *reviga* on at least two separate days, while all participants in the control group were retained for comparison. Missing data in the PP analyses were handled using the same multiple imputation approach as in the primary analysis.

ITT, J2R, and PP analyses were repeated at T2 to assess the durability of effects.

A responder analysis for the primary endpoint at T1 classified participants as responders if they showed an improvement of at least 6 points, corresponding to 15% of the scale range [6] (IQWiG, 2023). Proportions of responders were compared between groups using $\chi^2$ tests.

Throughout, a significance level of α = 0.05 (two-sided) was applied. To control for multiplicity, a gatekeeping strategy was pre-specified in the primary analysis, testing secondary endpoints in a predefined order, with each endpoint analyzed only if the previous one reached statistical significance [7].

### Supplementary Results

Supplementary Table 1 | Results of primary and secondary endpoints for J2R analyses

| | **Time** | **Control** | | | ***reviga*** | | | **ANCOVA** | | | |
|---|---|---|---|---|---|---|---|---|---|---|---|
| | | n | mean | SD | n | mean | SD | Treatment effect[a] (95% CI) | *p*-value | Partial $\eta^2$ | Cohen's *d* (95% CI)[b] |
| PSS-10 | T0 | 143 | 27.6 | 3.7 | 147 | 28.0 | 3.6 | - | - | - | - |
| | T1 | 143 | 24.0 | 5.8 | 147 | 22.5 | 6.4 | -1.8 (-3, -0.5) | 0.006 | 0.03 | 0.31 (0.09, 0.52) |
| | T2 | 143 | 23.3 | 6.4 | 147 | 21.1 | 6.8 | -2.4 (-3.7, -1.2) | < .001 | 0.04 | 0.39 (0.19, 0.59) |
| GAD-7 | T0 | 143 | 12.6 | 3.9 | 147 | 12.0 | 4.5 | - | - | - | - |
| | T1 | 143 | 10.2 | 4.9 | 147 | 9.0 | 4.5 | -1.0 (-1.8, -0.2) | 0.02 | 0.02 | 0.23 (0.04, 0.42) |
| | T2 | 143 | 10.1 | 4.9 | 147 | 8.4 | 4.8 | -1.4 (-2.2, -0.6) | 0.001 | 0.03 | 0.32 (0.13, 0.5) |
| WSAS | T0 | 143 | 24.6 | 6.9 | 147 | 24.6 | 6.8 | - | - | - | - |
| | T1 | 143 | 21.8 | 8.2 | 147 | 19.9 | 8.2 | -1.9 (-3.4, -0.4) | 0.013 | 0.02 | 0.26 (0.06, 0.47) |
| | T2 | 143 | 20.7 | 9.0 | 147 | 18.0 | 9.1 | -2.8 (-4.4, -1.2) | < .001 | 0.03 | 0.34 (0.15, 0.54) |
| OLBI | T0 | 143 | 2.9 | 0.4 | 147 | 2.9 | 0.3 | - | - | - | - |
| | T1 | 143 | 2.7 | 0.4 | 147 | 2.7 | 0.4 | -0.1 (-0.2, 0) | 0.013 | 0.02 | 0.26 (0.06, 0.46) |
| | T2 | 143 | 2.7 | 0.4 | 147 | 2.6 | 0.5 | -0.1 (-0.2, 0) | 0.005 | 0.02 | 0.28 (0.09, 0.47) |
| AQoL-8D | T0 | 143 | 56.2 | 9.3 | 147 | 55.0 | 8.9 | - | - | - | - |

| | | | | | | | | | | | |
|---|---|---|---|---|---|---|---|---|---|---|---|
| | T1 | 143 | 59.0 | 12.1 | 147 | 60.5 | 11.4 | 2.6 (0.8, 4.4) | 0.005 | 0.03 | 0.31 (0.09, 0.52) |
| | T2 | 143 | 60.0 | 12.9 | 147 | 62.1 | 12.6 | 3.3 (1.4, 5.2) | < .001 | 0.03 | 0.35 (0.16, 0.54) |
| Sick days | T0 | 143 | 12.3 | 21.3 | 147 | 14.3 | 23.9 | - | - | - | - |
| | T1 | 143 | 15.8 | 26.2 | 147 | 17.7 | 27.0 | 0.2 (-3.8, 4.3) | 0.906 | 0 | -0.18 (-0.33, -0.04) |
| | T2 | 143 | 13.5 | 24.5 | 147 | 13.0 | 24.3 | -1.6 (-5.8, 2.5) | 0.435 | 0 | -0.09 (-0.21, 0.03) |

[a] Group difference on original scale 3 months (T1) and 6 months (T2) after baseline, adjusted for baseline scores.

[b] based on baseline-adjusted means; positive values show effects in favor of the intervention group.
